# Determination of vancomycin exposure target and individualised dosing recommendations for neonates: model-informed precision dosing

**DOI:** 10.1101/2020.03.30.20045971

**Authors:** Zhe Tang, Jing Guan, Jingjing Li, Yanxia Yu, Miao Qian, Jiang Cao, Weiwei Shuai, Zheng Jiao

**Author notes:** Correspondence author Zheng Jiao, ORCID: 0000-0001-7999-7162, Department of Pharmacy, Shanghai Chest Hospital, Shanghai Jiao Tong University 241 Huai-hai West Road, 200030 Shanghai, China.

## Abstract

Few studies incorporating population pharmacokinetic/pharmacodynamic (Pop-PK/PD) modelling have been conducted to quantify the exposure target of vancomycin in neonates. To determine this target and dosing recommendations, a retrospective observational cohort study was established in neonates (chictr.org.cn, ChiCTR1900027919). A Pop-PK model was developed to estimate PK parameters. Causalities between acute kidney injury (AKI) occurrence and vancomycin use were verified using Naranjo criteria. Thresholds of vancomycin exposure in predicting AKI or efficacy were identified via classification and regression tree analysis. Associations between exposure thresholds and clinical outcomes including AKI and efficacy, were analysed by logistic regression. Dosing recommendations were designed using Monte Carlo (MC) simulations based on the optimised exposure target. Pop-PK modelling included 182 neonates with 411 observations. In covariate analysis, neonatal physiological maturation, renal function, and concomitant use of vasoactive drugs (VAS) significantly affected vancomycin pharmacokinetics. Seven cases of vancomycin-induced AKI were detected. Area under the concentration–time curve from 0–24 h (AUC_0-24_) ≥ 485 mg·h/L was an independent risk factor for AKI after adjusting for VAS co-administration. Clinical efficacy of vancomycin was analysed in 42 patients with blood culture-proven staphylococcal sepsis. AUC_0–24_ to minimum inhibitory concentration (AUC_0–24_/MIC) ≥ 234 was the only significant predictor of clinical effectiveness. MC simulations indicated that regimens in Neonatal Formulary 7 and Red Book (2018) were not suitable for all neonates. In summary, AUC_0-24_ of 240-480 assuming MIC = 1 mg/L is a recommended exposure target of vancomycin in neonates. Model-informed dosing regimens are valuable in clinical practice.

**Highlights:**

- The optimised neonatal exposure target of vancomycin was uniquely quantified
- Day 1 exposure of vancomycin is the predictor for clinical outcomes in neonates
- Vancomycin-induced acute kidney injury is not related to high exposure alone
- A lower exposure target is likely to be more effective for neonates than for adults
- Monte Carlo simulation provides a more suitable dosing regimen for neonates

## 1. Introduction

Neonates are predisposed to bacterial sepsis owing to their compromised immunity. *Staphylococcus aureus* and other *Staphylococcus* spp. are the predominant pathogenic bacteria[1]. Vancomycin is the first-line choice against staphylococcal sepsis for neonates in intensive care units (ICUs).

The recent consensus guideline[2] supports a Bayesian-derived area under the concentration–time curve to minimum inhibitory concentration (AUC/MIC) of 400 to 600 (assuming MIC=1 mg/L) as the vancomycin exposure target for adults infected by methicillin-resistant *S. aureus* (MRSA). Bayesian-derived AUC monitoring is advocated, whereas trough target-based dosing is not recommended, given that an identical trough value can be derived from a wide range of daily AUC values[2, 3].

Several published reports on adults have examined vancomycin exposure-outcome relationships by incorporating population pharmacokinetic/pharmacodynamic (Pop-PK/PD) modelling[4-9]. Comparatively, few exposure-outcome evaluations have been conducted in neonates[10]. Vancomycin is known as a potential nephrotoxin. However, its exposure threshold associated with vancomycin-induced acute kidney injury (AKI) has not been clearly defined in neonates[10]. The association between vancomycin exposure and clinical efficacy has not been clarified in this population either[11]. Thus, there is a critical need for clinical studies in neonates that associate individual exposure with clinical outcomes incorporating AKI and efficacy.

The aim of this study was to: (i) derive vancomycin exposure target that balances both AKI and efficacy in ICU neonates, with a focus on AUC/MIC; (ii) optimise neonatal dosing recommendations of vancomycin using Monte Carlo (MC) simulation according to the exposure target obtained and the established Pop-PK model.

## 2. Patients and methods

### 2.1. Study population

A retrospective observational cohort study was conducted and executed in accordance with the recommendations of the Declaration of Helsinki (2000). The protocol was approved by the Ethics Committee of Nanjing Maternity and Child Health Care Hospital. The identities of patients were hidden during the study. Informed consent was approved unnecessary owing to the retrospective nature of the study. This study was registered at www.chictr.org.cn (ChiCTR1900027919).

The electronic medical records of patients who suffered from suspected or confirmed neonatal sepsis and received vancomycin treatment in the neonatal ICU from the 1^st^ of January 2016 to the 31^st^ of December 2019 were reviewed to determine eligibility. Clinical diagnosis of neonatal sepsis followed the criterion by Auriti et al[12]. Patients with postmenstrual age (PMA)≤48 weeks when vancomycin was initiated were included. All individuals had normal baseline serum creatinine (SCR) as defined by both gestational age (GA) and postnatal age (PNA) at the start of vancomycin treatment[13, 14]. They were treated with vancomycin for ≥3 days, with at least one vancomycin serum concentration being monitored. The exclusion criteria included vancomycin serum concentrations lower than the limit of detection, a diagnosis of congenital anomalies of the kidney and urinary tract, major congenital heart disease, or congenital haematological malignancy, and experience of AKI prior to vancomycin treatment.

Information of interest included: demographic information, dosing history, vancomycin serum concentration, SCR, creatinine clearance (CLCR), co-medications, clinical diagnosis, radiographic examinations, and microbiological culture. CLCR was calculated by the Schwartz equation[15] (Eq. 1).

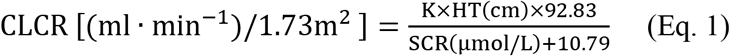

Where HT is body length of neonates, SCR is serum creatinine, and K is a constant equal to 0.33 (preterm neonates with PMA < 37 weeks) or 0.45 (term neonates with PMA ≥ 37 weeks).

According to the local protocol, neonates with suspected or confirmed sepsis received empirical therapy with vancomycin in combination with meropenem based on the presumptive diagnosis. Once a diagnosis was established, anti-infectives were rationalised to cover the causative pathogen.

Vancomycin (10-15 mg/kg; Vancocin, Lilly, S.A, Suzhou, China) was administered every 8 to 18 h with intermittent infusion lasting 60 min. Blood samples were collected 0.5 h before vancomycin administration (trough) or 0.5 h after infusion completion (peak) prior to the fourth or fifth dose. Target trough levels were 10 to 20 mg/L[16].

### 2.2. Bioanalytical method

Vancomycin concentrations were analysed by a fluorescence polarisation immunoassay using the ARCHITECT i2000SR (Abbott Laboratories, Chicago, IL, USA) within 24 h of sample collection. The limit of detection was 1 mg/L, and the calibration range was 3 to 100 mg/L. The intra-day and inter-day coefficients of variation (CV) were less than 20%. SCR was analysed using an enzymatic method (Beckman Coulter AU5800, Beckman Coulter Inc., Brea CA, USA) with linearity from 5 to 13275 μmol/L. The intra-day and inter-day CVs were less than 5%.

Etiological examinations were provided by the clinical laboratory of our hospital using the VITEK 2 microbial identification system (bioMérieux, Lyon, France). The broth microdilution (BMD) method was used to determine MICs. The interpretative reports were issued by laboratory technicians according to the criteria of The Clinical and Laboratory Standards Institute (CLSI)[17].

### 2.3. Pop-PK modelling

A Pop-PK model was developed to estimate PK parameters using nonlinear mixed effect modelling software Monolix (version 2019R2, Lixoft, France) with the Stochastic Approximation Expectation Maximization (SAEM) algorithm.

A one or two compartment model with first-order elimination was examined to fit the dataset. Interindividual variabilities in PK parameters were estimated using an exponential model. Residual unexplained variabilities were tested by an additive model, an exponential model, or a combined additive and exponential model. Potential covariates were screened, and concomitant medication present in less than 20% of the population was not tested.

Five models were employed to describe the impacts of size (weight) and maturation (age) on clearance (CL) (Table S1)[18–20]. The model with the lowest Akaike information criteria (AIC) and Bayesian information criteria (BIC), as well as appropriate estimates of parameters, was employed for further analysis[21]. The selection of covariates was determined using a forward inclusion and a backward elimination process. Nested covariate models were statistically compared using a likelihood ratio test on differences in the objective function value (OFV). A reduction in OFV of 3.84 (P < 0.05) for forward inclusion and an increase in OFV of 6.63 (P < 0.01) for backward elimination were the criteria for retaining a covariate in the model.

The final model was internally assessed by visual inspection of goodness-of-fit plots, normalised prediction distribution error (NPDE), and bootstrap[21-23]. The data from a previous study by Li et al.[24] were used to validate the final model externally. The mean prediction error (MPE) and mean absolute prediction error (MAPE) (Table S2) were applied to calculate the bias and imprecision of model predictions[25].

### 2.4. Estimation of individual exposure variables

Individual exposure variables for the initial 48 h of vancomycin therapy, including day 1 AUC (AUC_0-24_), day 2 AUC (AUC_24-48_), and cumulative day 1 and 2 AUC (AUC_0-48_), were estimated by Simulx (version 2019R2, Lixoft, France). Although there were no data on which AUC (i.e., initial AUC or AUC at steady state (AUC_ss_)) to use in determining the vancomycin exposure target, exposure to initial therapy may be more appropriate because the potential influence of the worsening of infection on vancomycin CL could be avoided. Further, the AUC in the first 48 h is critical for ICU neonates and has been associated with clinical outcomes in adults[9, 26].

### 2.5. Determination of the exposure target

#### 2.5.1. AKI analysis

All patients were screened for AKI during vancomycin treatment. The definition of AKI included an increase in SCR of ≥ 26.5 μmol/L within 48 h, or ≥ 1.5 times from the previous lowest value within 7 days, or a urine volume < 1.0 mL/(kg·h)[27]. To identify whether the occurrence of AKI was related to the use of vancomycin, causality analyses were performed using the Naranjo criteria[28]. AKIs classified as definite, probable or possible by the Naranjo criteria, were considered to be vancomycin-induced side effects, and were eligible for further analyses. AKIs classified as doubtful were excluded from the study.

#### 2.5.2. Efficacy analysis

The clinical efficacy was analysed for patients with blood culture-proven staphylococcal sepsis. Exclusion criteria were as follows: vancomycin treatment ≤ 5 days, polymicrobial infection or concomitant use of an antibiotic sensitive to the identified staphylococcus. Vancomycin treatment failure was defined as positive blood culture ≥ 5 days after the initiation of treatment, recurrence of bacteraemia within 30 days, or 30-day mortality[5, 7, 26].

Thresholds of vancomycin exposure in predicting AKI or efficacy were derived using classification and regression tree (CART) analysis (R package rpart, version 4.1-15). The predictive performance of thresholds, comprising CART-derived and other priori-defined ones, were evaluated using receiver operating characteristic (ROC) curves, along with predictive matrices including negative and positive predictive values (NPV and PPV, respectively) (R package pROC, version 1.16.2)[9, 29]. Logistic regression analyses were performed to quantify associations between vancomycin exposure and clinical outcomes. Other potential confounding variables with P < 0.20 in the univariate analysis were included in multivariate analyses using a stepwise approach and retained in the model. If multiple variables were correlated, only one was tested. P values of ≤ 0.05 were considered statistically significant.

According to the analyses of AKI and efficacy, the optimised exposure target was set as the basis for the following dosing regimen design.

### 2.6. Model-informed dosing regimen

Dosing regimens were designed and compared with the recommendations in Neonatal Formulary 7[30] and Red Book (2018)[31]. Virtual patients with different physiological characteristics were employed for MC simulations. Demographic information was obtained from the Fenton growth chart[32]. A total of 1000 replicates for each scenario were simulated to detect the most proper scheme (R package mlxR, version 2.0.2).

## 3. Results

### 3.1. Study population

One hundred and eighty-five neonates diagnosed with suspected or confirmed neonatal sepsis received intermittent intravenous infusion of vancomycin in our institution from the 1^st^ of January 2016 to the 31^st^ of December 2019. There were 182 patients eligible for this study, with 286 trough and 125 peak concentrations (Table S3). Two patients with incomplete data and one patient diagnosed with haematological malignancy were excluded (Fig. 1).

**Fig. 1.**
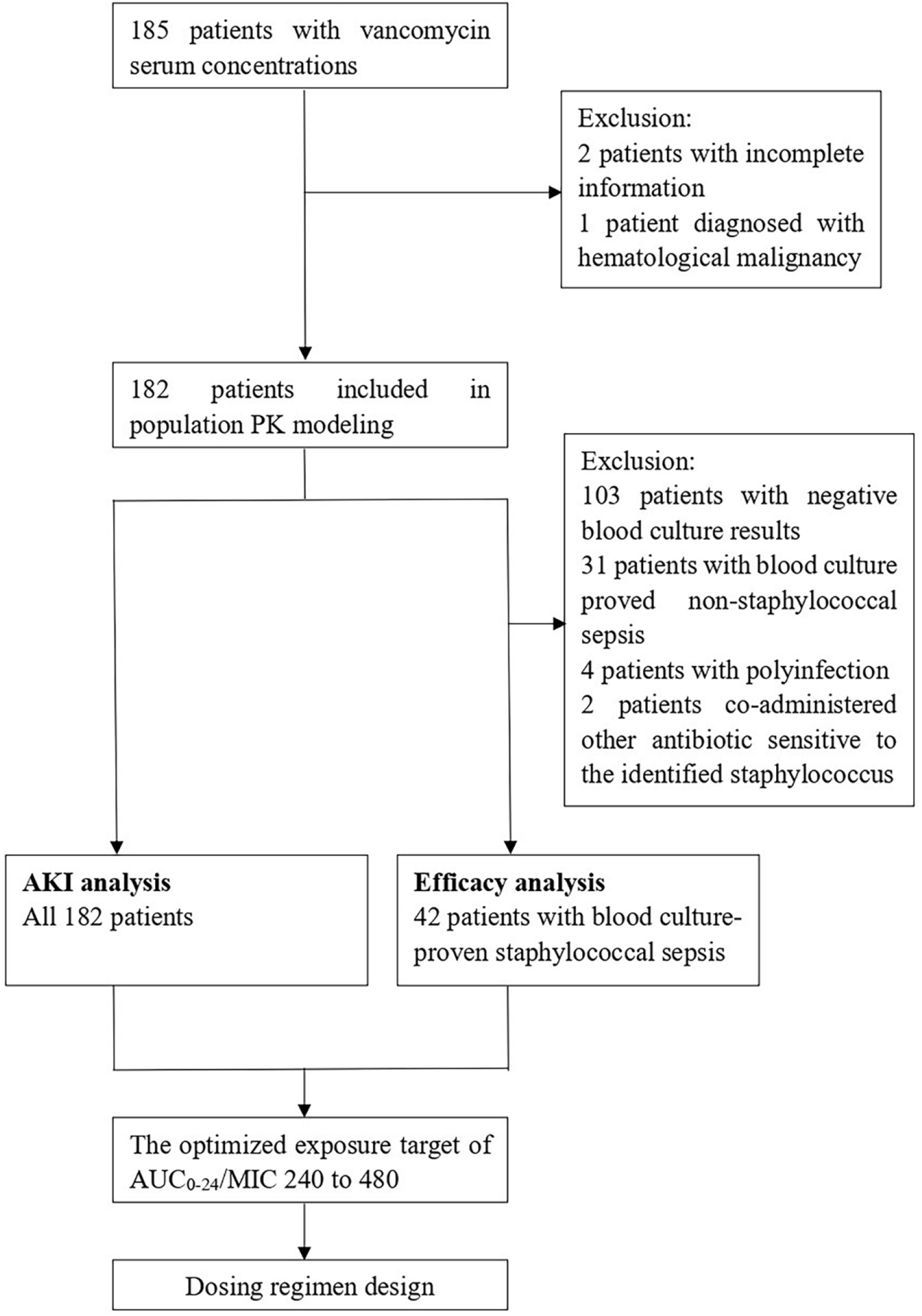
Study flow.

### 3.2. Pop-PK modelling

A Pop-PK model was developed to estimate individual parameters. A one compartment model with first-order elimination was sufficient to fit the data. A proportional model was chosen to describe residual unexplained variabilities. The detailed modelling process is summarised in Table S4.

Both, Model II with PMA, and Model III had relatively lower AIC and BIC. Model II with PMA was selected because of its simplicity. SCR, CLCR, and the co-administration of vasoactive drugs (VAS) were recognised as significant covariates on CL. As SCR and CLCR were correlated and performed equally, SCR was retained owing to better clinical accessibility. A 14% decrease in CL was associated with the co-administration of VAS. Moreover, current weight (cWT) was incorporated into the volume of distribution (V) for physiological plausibility. The results of the final model are listed in Table 1.

**Table 1.** The estimates of the final model and 2000 bootstrap results.

| Parameter<sup>a</sup> | Final model |  | Bootstrap |  | Bias (%)<sup>c</sup> |
| --- | --- | --- | --- | --- | --- |
|  | Estimate | RSE (%)<sup>b</sup> | Median | 2.5%–97.5% |  |
| **Structure model parameter** |  |  |  |  |  |
| V<sub>pop</sub> (L) | 0.86 | 3.02 | 0.87 | 0.80 – 0.93 | 0.82 |
| CL<sub>pop</sub> (L/h) | 0.09 | 2.11 | 0.09 | 0.08 – 0.09 | 0.69 |
| beta1 | 2.35 | 1.20 | 2.29 | 1.94 – 2.66 | –2.77 |
| beta2 | 0.18 | 3.41 | 0.18 | 0.09 – 0.28 | 4.55 |
| F<sub>VAS</sub> | 0.86 | 0.34 | 0.85 | 0.77 – 0.94 | –1.28 |
| **Inter-individual variability** |  |  |  |  |  |
| V (%CV) | 11% | 24.20 | 14% | 0.09 – 0.20 | 24.11 |
| CL (%CV) | 24% | 6.85 | 23% | 0.20 – 0.26 | –2.11 |
| **Residual variability** |  |  |  |  |  |
| Proportional (%CV) | 0.21 | 4.82 | 0.21 | 0.18 – 0.24 | –0.47 |

V<sub>pop</sub>, population mean estimate for volume of distribution in a subject with a body weight of 1.5 kg; cWT, current body weight; CL<sub>pop</sub>, population mean estimate for vancomycin clearance in a subject with a body weight of 1.5 kg; PMA, postmenstrual age; SCR, serum creatinine; F<sub>VAS</sub>, scaling factor of concomitant use of vasoactive drugs.
<sup>a</sup>V (%CV), interindividual variability of volume of distribution; CL (%CV), interindividual variability of vancomycin clearance.
<sup>b</sup>RSE, relative standard error.
<sup>c</sup>Bias%, relative bias of estimates by Monolix to the median estimates by 2000 bootstrap procedures. The rate of bias in fixed effects less than 5% was thought acceptable. It was calculated by the formula $Bias(\%) = (Median_{bootstrap} - Median_{final\ model}) / Median_{final\ model} \times 100\%$ .

The goodness-of-fit plot of the final model showed no obvious bias or trend (Fig. 2). The results of NPDE generally satisfied the hypothesis of normal distribution (Fig. 3). Bootstrap analysis (n = 2000) showed that the final model was precise, because the median parameter estimates were close to those of the final model (Table 1).

**Fig. 2.**
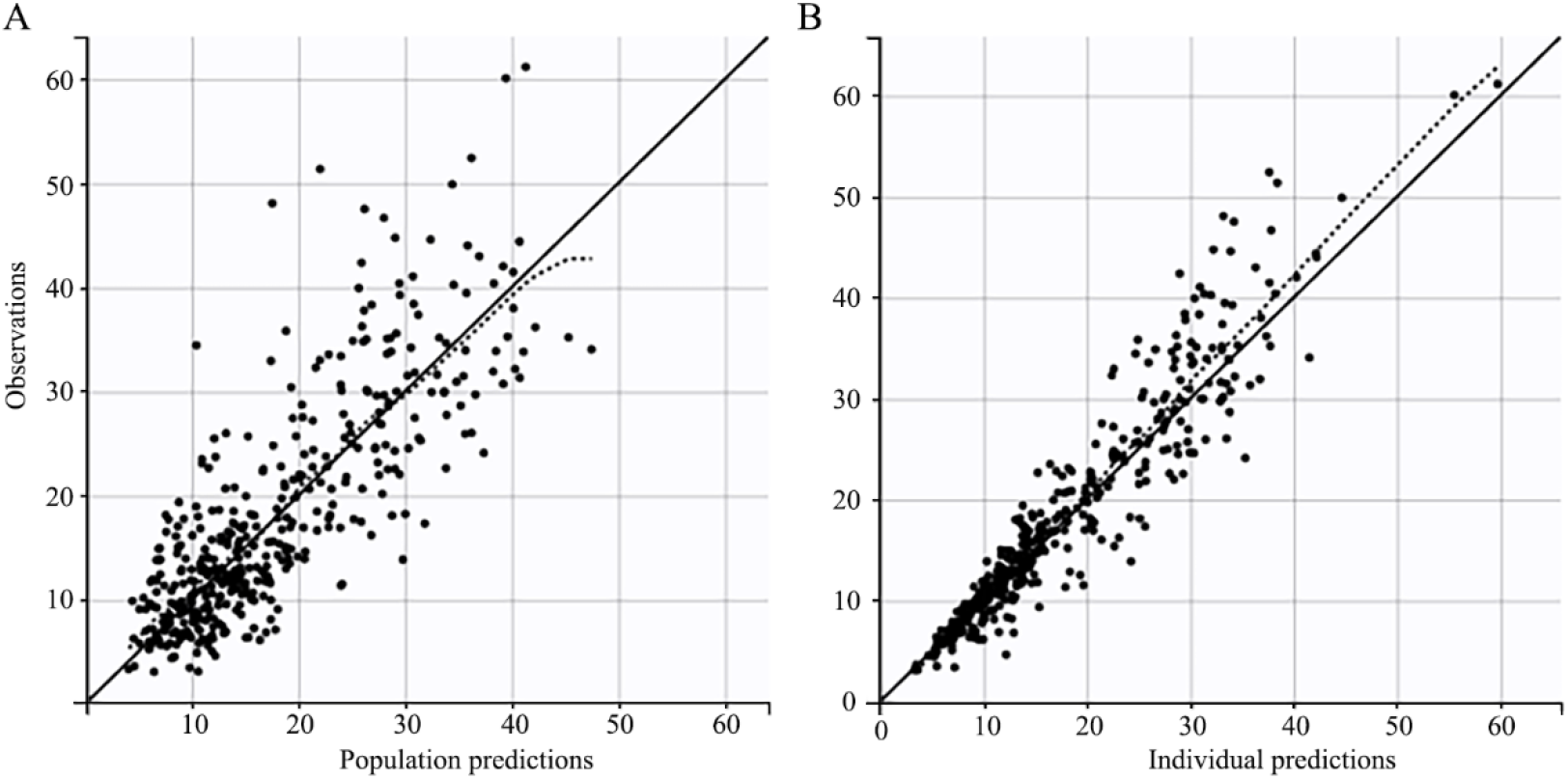
Goodness-of-fit plot of the final established Pop-PK model. (A) Population predictions vs. observations. (B) Individual predictions (IPRED) vs. observations. The black solid lines are the reference lines, and black dashed lines are the loess smooth lines.

**Fig. 3.**
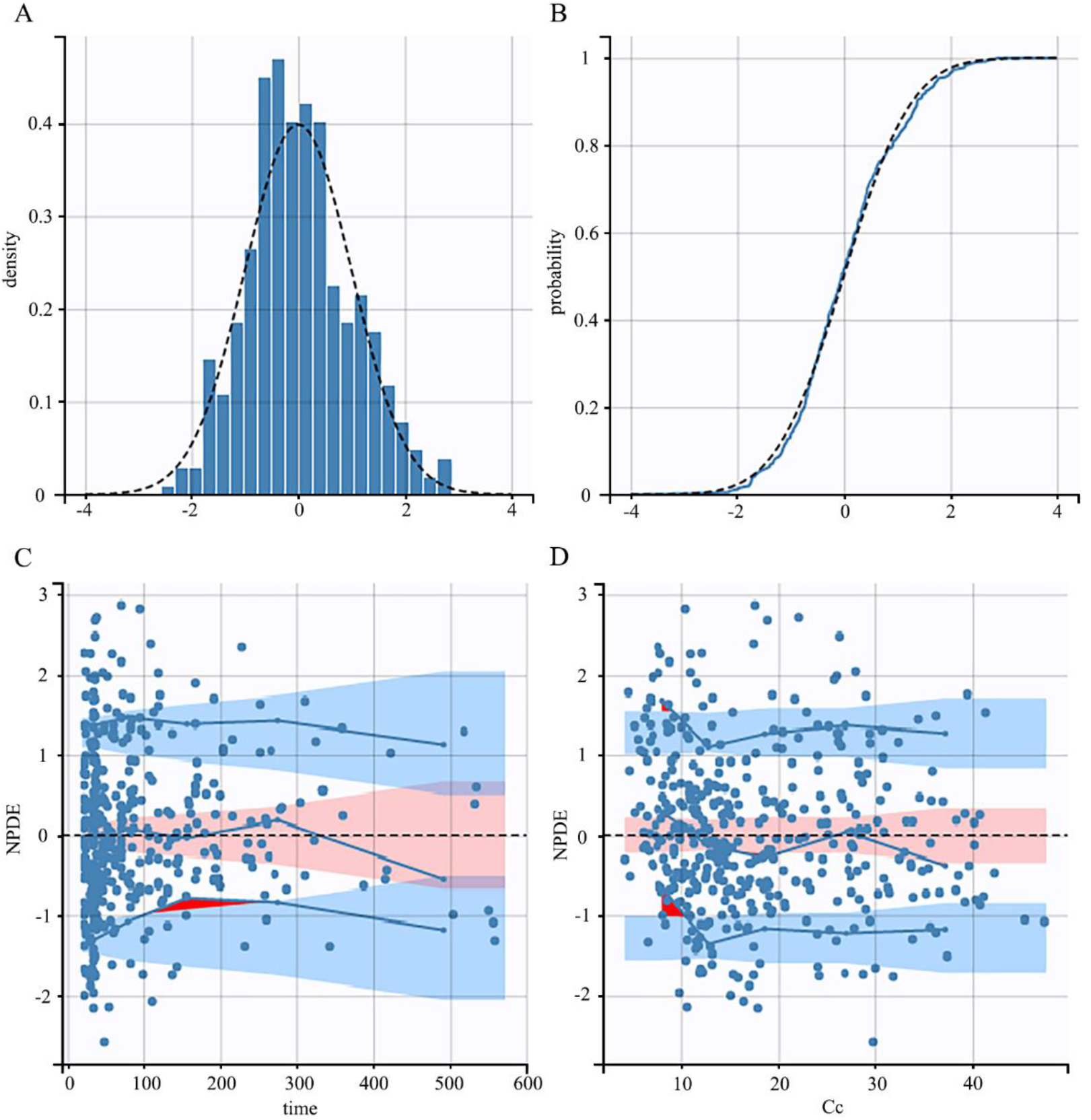
Normalized prediction distribution error (NPDE) plots for the final model. (A) Histogram of NPDE values; (B) NPDE vs. the probability density. Black dashed lines, theoretical distribution. Blue dashed line, empirical distribution. (C) Scatter plot of the time vs. NPDE; (D) Scatterplot of serum concentrations (Cc) vs. NPDE. Blue filled dots, observed data; Black dashed lines, theoretical mean; Blue solid lines, empirical percentiles of observations; Blue-shaded area, 95% confidence interval (CI) for the 5th and 95th percentiles of the predicted values. Pink-shaded area, 95% CI for the 50th percentiles of the predicted values; Red-shaded area, outliers.

In the external evaluation, 28 patients in an earlier study by Li et al.[24] were included. All observations were troughs. The medians for cWT, PMA, and SCR were 1.66 kg (range, 0.80 to 2.52 kg), 33.22 weeks (range, 28.28 to 36.71 weeks), and 35.42 μmol/L (range, 14.17 to 51.71 μmol/L), respectively. No one was co-medicated with VAS. The MPE and MAPE for the final model were 3.97% (standard deviation, 19.88%) and 13.01% (standard deviation, 15.36%), respectively.

As the external evaluation showed no obvious bias, we combined all data to re-estimate the PK parameters. The estimates did not show an apparent difference (Table S5).

### 3.3. Determination of the exposure target

#### 3.3.1 AKI analysis

Seven AKI cases were identified in total. Five occurred on day 3 after vancomycin was initiated, and two on day 4. Patients who experienced AKI had higher baseline SCR and lower CLCR than those who did not experience AKI. Although there was no statistical difference in daily dose between patients with and without AKI, patients with AKI had significantly higher vancomycin exposure quantified by AUC_0-24_, AUC_24-48_, and AUC_0-48_ owing to their decreased vancomycin CL. (Table 2).

**Table 2.**
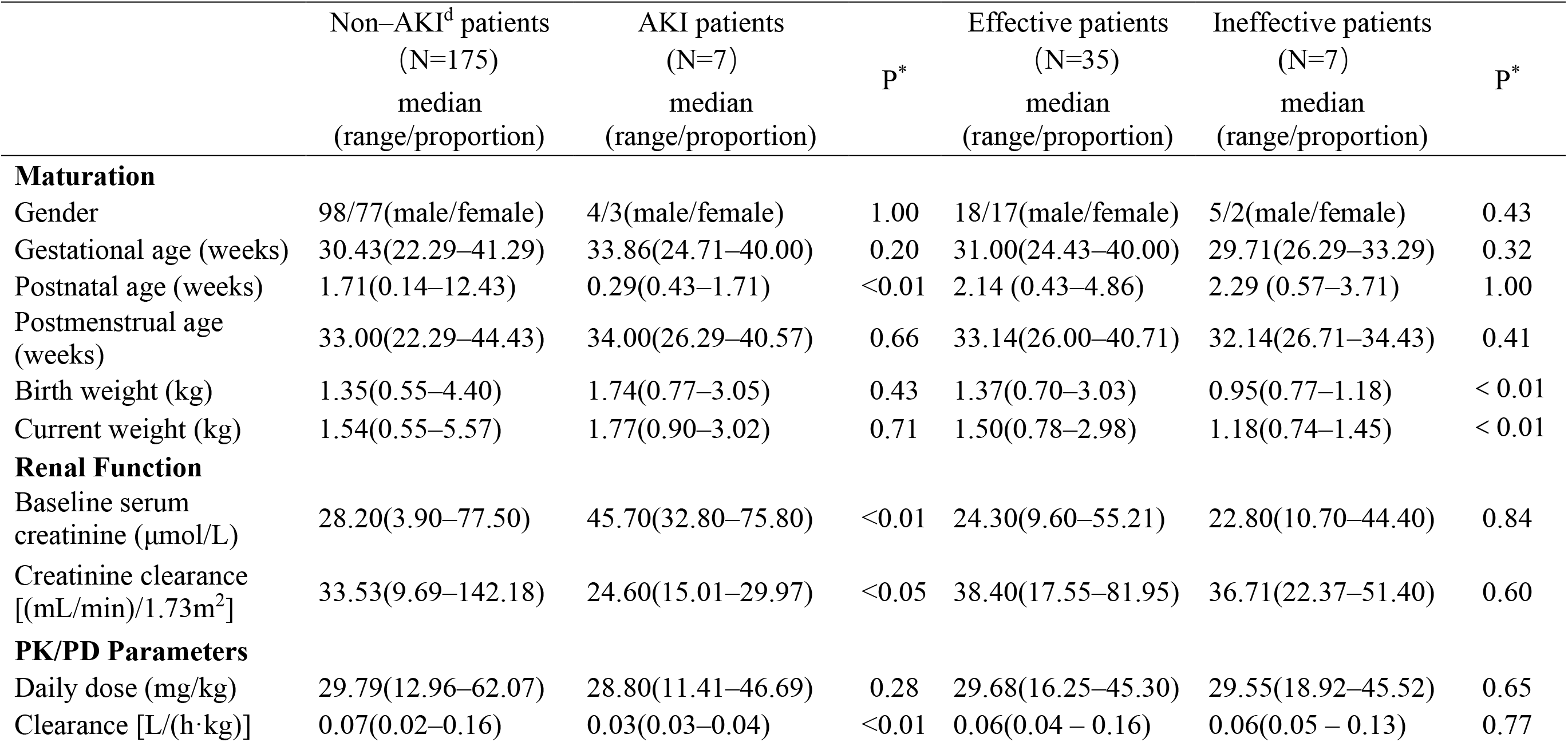
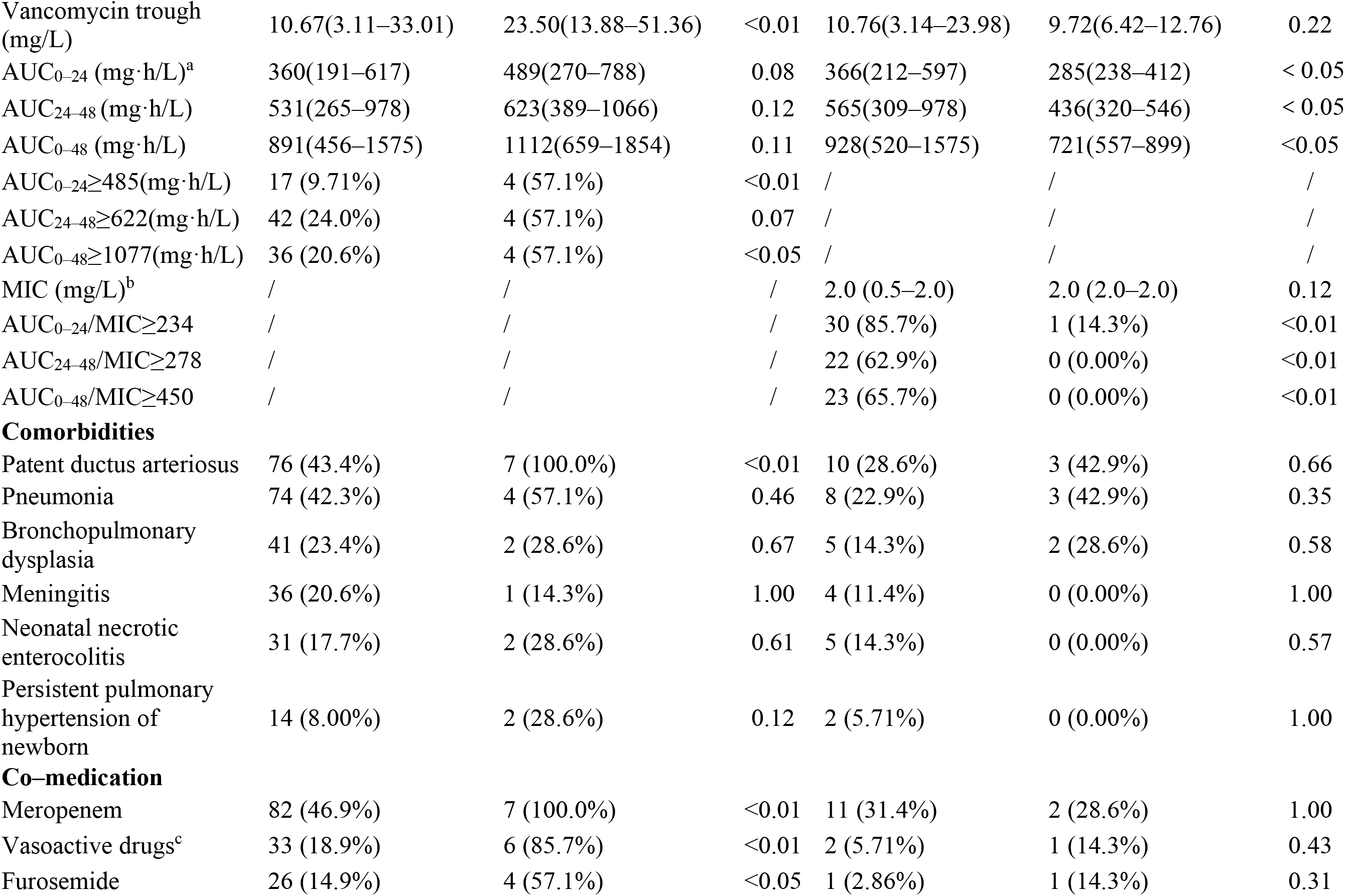
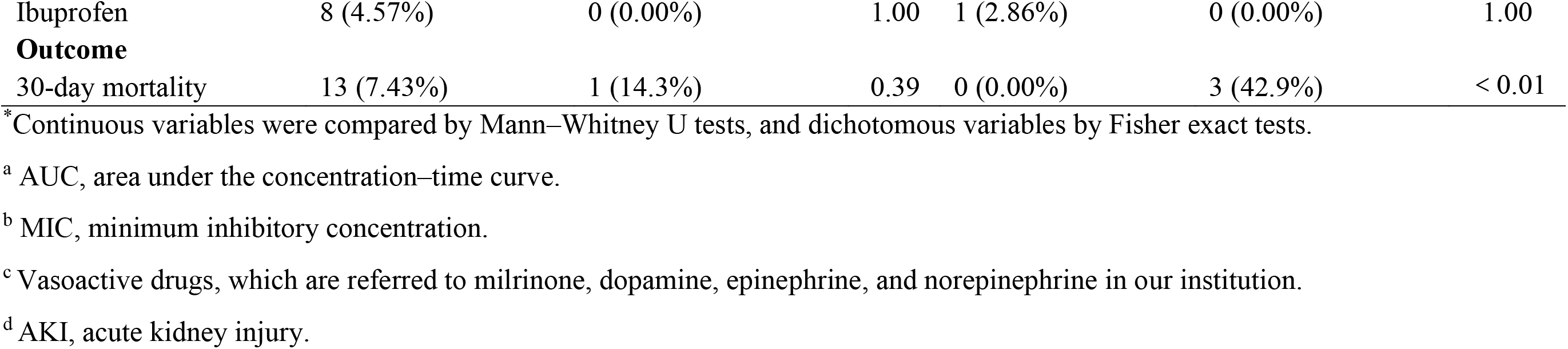
Comparisons of demographic and PK/PD characteristics between patients with and without AKI / patients with and without effective treatment.

With regard to the causality between AKI occurrence and the use of vancomycin, causalities for four cases were classified as probable, and three cases as possible using the Naranjo criteria. Hence, all AKI cases were eligible for the following analyses. Vancomycin dosages were immediately reduced for six patients after AKI occurrences. Then their AKI symptoms were alleviated, and renal function recovered within 5 days.

VAS, prescribed to 39 patients, was the major potential nephrotoxin besides vancomycin. VAS refers to milrinone, dopamine, epinephrine, and norepinephrine in our institution. Six of seven AKI cases were co-administered VAS.

In ROC curve analysis, AUC_0-24_, AUC_24-48_, and AUC_0-48_ significantly predicted AKI with good performance (Fig. 4A). The CART-derived threshold of AUC_0-24_ ≥ 485 mg·h/L had the highest predictive value with an area under the ROC curve (AUCROC) of 0.74 (confidence interval (CI): 0.54, 0.94) (Table S6). PPV, which represented true-positive predictions of patients who experienced AKI, was positively correlated with vancomycin exposure. At AUC_0-24_≥485 mg·h/L, PPV increased more rapidly, indicating an additional risk of AKI (Table S6). Therefore, this threshold was included in further multivariate regression analyses.

**Fig. 4.**
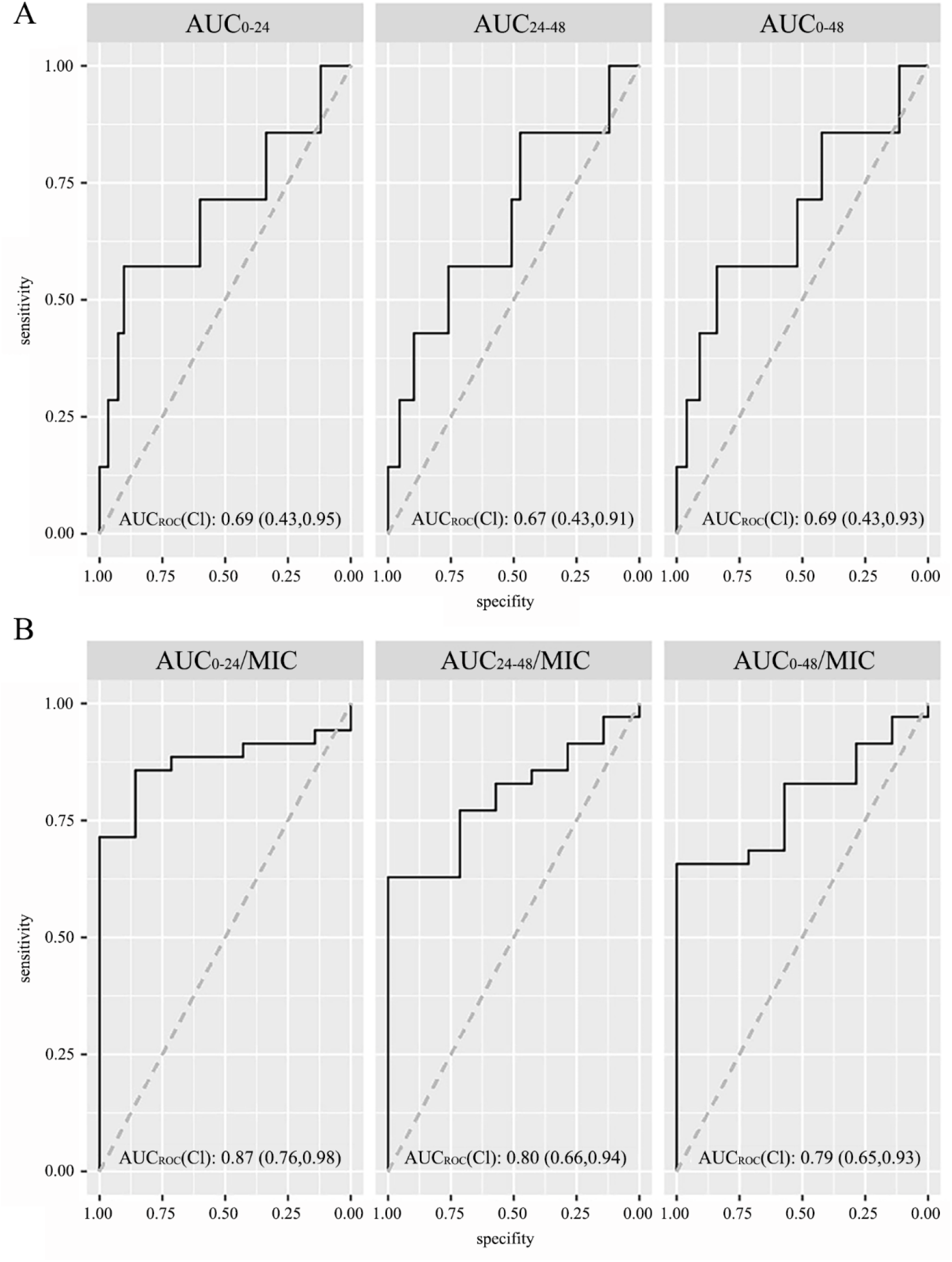
Receiver operating characteristic (ROC) curves. (A) ROC curves of AUC_0-24_, AUC_24-48_ and AUC_0-48_ for prediction of AKI. (B) ROC curves of AUC_0-24_/MIC, AUC_24-48_/MIC and AUC_0-48_/MIC for prediction of efficacy. AUC, area under the concentration-time curve; MIC, minimum inhibitory concentration; AUCroc, area under the ROC curve.

In univariate regression analysis, PNA, persistent pulmonary hypertension of newborn, and concomitant use of VAS or furosemide were identified as potential confounding factors that influenced AKI besides AUC_0-24_ (Table S7). The vancomycin trough was not included in multivariate analysis because it was strongly correlated with AUC_0-24_ (Spearman coefficient = 0.75, P < 0.01). SCR and CLCR were also excluded as they were included in the diagnostic criteria for AKI. In multivariate regression analysis, AUC_0-24_ ≥ 485 mg·h/L was independently associated with AKI after adjusting for the concomitant use of VAS (Table 3).

**Table 3.**
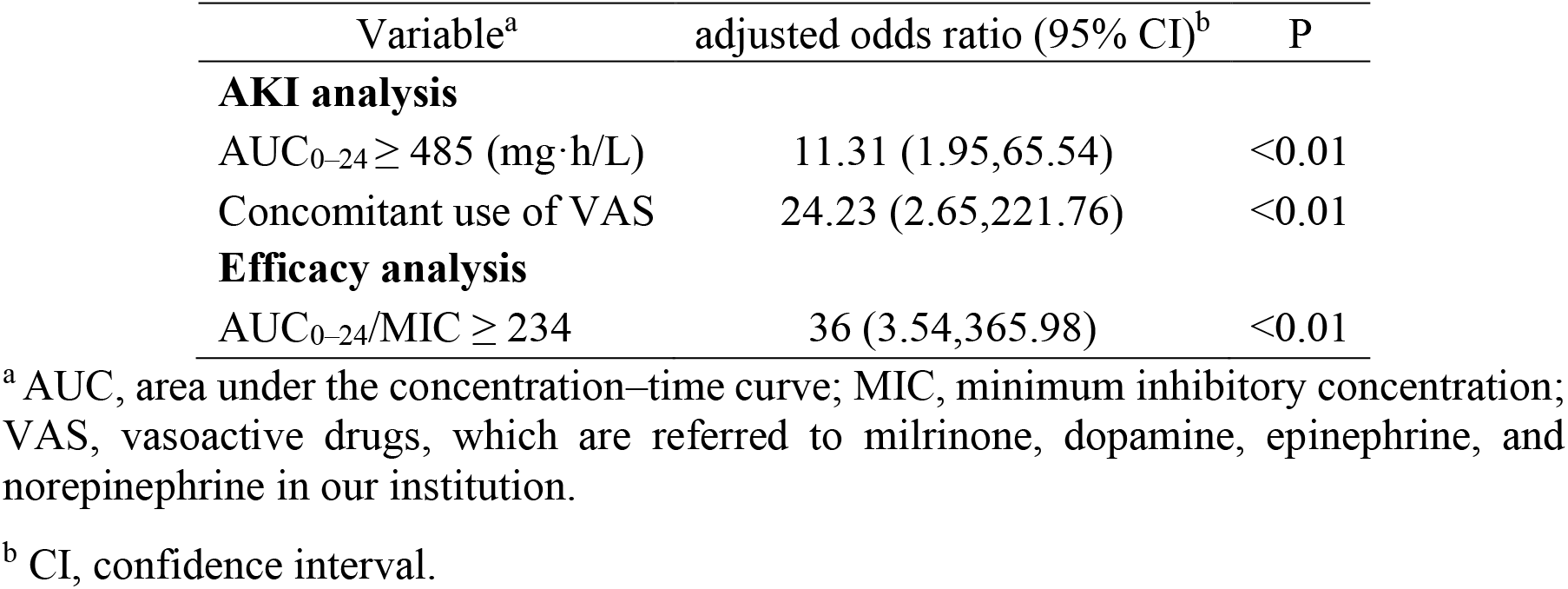
Results of multivariate regression analysis.

#### 3.3.2 Efficacy analysis

Seventy-nine patients had positive blood cultures. Thirty-one with blood culture-proven non-staphylococcal sepsis, four with polymicrobial infections, and two with co-administration of an antimicrobial that affected the identified staphylococcal species were excluded. Forty-two patients with blood culture-proven staphylococcal sepsis were eligible for efficacy analysis (Fig. 1). Sixteen of them were infected by *S. aureus*, whereas the others were infected by other *Staphylococcus* spp.

Seven patients experienced treatment failures. Three died during vancomycin treatment. Four had positive blood culture results ≥ 5 days after the initiation of vancomycin, and linezolid was used as an alternative. No one experienced a recurrence of bacteraemia. Despite the insignificant difference in daily dosage and MICs between patients with and without effective treatment, vancomycin exposures were much lower among ineffective patients quantified by AUC_0-24_/MIC, AUC_24-48_/MIC, and AUC_0-48_/MIC (Table 2).

ROC analyses showed that AUC_0-24_/MIC, AUC_24-48_/MIC and AUC_0-48_/MIC performed well in the prediction of efficacy (Fig. 4B). The threshold of AUC_0-24_/MIC ≥ 234 had the highest predictive value with an AUC_roc_ of 0.86 (CI: 0.71, 1.00) (Table S8). NPV, which referred to true-negative predictions of patients who experienced treatment failure, was negatively correlated with vancomycin exposure. At AUC_0-24_/MIC ≥ 234, PPV remained low, indicating a decreased risk of treatment failure (Table S8), which is why this threshold was included in further analyses.

Birth weight and cWT were identified as potential confounding factors that influenced efficacy, besides AUC_0-24_/MIC, by univariate regression analysis (Table S7). These two variables were apparently correlated. cWT was included in multivariate analyses because it better reflected the actual physiological status during vancomycin treatment. In multivariate regression analysis, AUC_0-24_/MIC ≥ 234 was the only significant factor associated with efficacy (Table 3).

MIC distributions from The European Committee on Antimicrobial Susceptibility Testing (EUCAST) show that 75% of *S. aureus* isolates and 45% of other *Staphylococcus* spp. isolates have a MIC of 1 mg/L to vancomycin[33]. According to analyses above, when MIC > 1 mg/L, the probability of achieving the exposure threshold for efficacy (AUC_0-24_/MIC ≥ 234) was low as the exposure was lower than the threshold for AKI (AUC_0-24_ ≥ 485). Therefore, the optimised exposure target for vancomycin in neonates was set as an AUC_0-24_ of 240 to 480, assuming MIC = 1 mg/L.

### 3.4. Model-informed dosing regimen

The dosage recommendations by the references[30, 31] and this study are presented in Table S9 and Table 4 respectively.

**Table 4.** Results of model–informed dosing regimens.

| cWT<sup>a</sup> (kg) | PMA<sup>b</sup> (weeks) | SCR<sup>c</sup> (μmol/L) | Patients without VAS<sup>d</sup> (mg/kg) | Patients with VAS (mg/kg) |
| --- | --- | --- | --- | --- |
| 0.7–0.99 | 25–27 | 10 | 17.5–20, q24h | 15–17.5, q24h |
|  |  | 25 | 15–17.5, q24h | 15–17.5, q24h |
|  |  | 45 | 15–17.5, q24h | 12.5–15, q24h |
|  |  | 60 | 15–17.5, q24h | 12.5–15, q24h |
|  |  | 90 | 12.5–15, q24h | 12.5–15, q24h |
| 1.0–1.69 | 28–31 | 10 | 15–17.5, q18h | 17.5–20, q24h |
|  |  | 25 | 12.5–15, q18h | 17.5–20, q24h |
|  |  | 45 | 12.5–15, q18h | 15–17.5, q24h |
|  |  | 60 | 12.5–15, q18h | 15–17.5, q24h |
|  |  | 90 | 10–12.5, q18h | 15–17.5, q24h |
| 1.7–2.69 | 32–36 | 10 | 15–17.5, q12h | 12.5–15, q12h |
|  |  | 25 | 12.5–15, q12h | 10–12.5, q12h |
|  |  | 45 | 10–12.5, q12h | 10–12.5, q12h |
|  |  | 60 | 10–12.5 q12h | 10–12.5, q12h |
|  |  | 90 | 10–12.5, q12h | 10, q12h |
| 2.7–3.69 | 37–41 | 10 | 17.5–20, q12h | 15–17.5, q12h |
|  |  | 25 | 15–17.5, q12h | 12.5–15, q12h |
|  |  | 45 | 12.5–15, q12h | 12.5–15, q12h |
|  |  | 60 | 12.5–15, q12h | 12.5–15, q12h |
|  |  | 90 | 12.5–15, q12h | 10–12.5, q12h |
| 3.7 – 4.6 | 42–46 | 10 | 12.5–15, q8h | 17.5–20, q12h |
|  |  | 25 | 12.5–15, q8h | 15–17.5, q12h |
|  |  | 45 | 10–12.5, q8h | 15, q12h |
|  |  | 60 | 10–12.5, q8h | 15, q12h |
|  |  | 90 | 10, q8h | 12.5–15, q12h |

<sup>a</sup> cWT, current body weight.
<sup>b</sup> PMA, postmenstrual age.
<sup>c</sup> SCR, serum creatinine.
<sup>d</sup> VAS, vasoactive drugs, which are referred to milrinone, dopamine, epinephrine, and norepinephrine in our institution.

Generally, patients with concomitant use of VAS requires a reduced daily dose. The dosing interval of 24 h is suitable for extremely preterm patients (PMA < 28 weeks), with a dose of 12.5-20 mg/kg. For preterm patients with a PMA between 28 and 36 weeks, proper dosing intervals varies from 12 to 24 h. Term neonates (PMA ≥ 37 weeks) require dosing intervals between 8 and 12 h. Dosing intervals ≤ 8 h are not necessary.

Our results suggest that the daily dose for extremely-low-birth-weight (ELBW) neonates (cWT < 1 kg) is 12.5 to 20 mg/kg (Table 4). However, Neonatal Formulary 7[30] recommends a daily dose of 20 mg/kg for all ELBW neonates. The maximum daily dose recommended by Red Book (2018)[31] for these patients is 30 mg/kg. Quite a few of ELBW neonates are overdosed in these references. Table 4 also shows that a daily dose of 20 to 45 mg/kg is required for normal-birth-weight (NBW) neonates with cWT > 2.7 kg and SCR ≤ 60 μmol/L, whereas Red Book (2018)[31] suggests a daily dose of 30 mg/kg for patients (GA > 28 week) with normal renal function (SCR ≤ 61.7 μmol/L). Apparently, this regimen is unable to satisfy the treatment needs of all NBW neonates.

## 4. Discussion

To the best of our knowledge, this is the first study in neonates uniquely utilising Pop-PK/PD modelling with Bayesian estimation to evaluate vancomycin exposure associated with clinical outcomes including AKI and efficacy. With Bayesian estimation, PK/PD variability for vancomycin exposure-outcomes evaluations can be plausibly quantified. Bayesian estimation also allows for more accurate and reliable quantification of exposure parameters (i.e. AUC) with sparse data. Moreover, using Pop-PK based on MC simulations, dosing recommendations for neonates can be derived based on the optimised exposure target.

The exposure threshold for vancomycin-induced AKI in neonates has been clearly defined in this study for the first time. The threshold for AKI, AUC_0-24_ of 480 mg·h/L, is lower than that in previous report on children. Le et al.[34] suggests that AUC_ss_ over 24 h in children should be less than 800 mg·h/L to minimise AKI risk. The time to AKI in Le’s study ranged from 1 to 15 days[34]. The reported threshold based on AUC_ss_ is probably inapplicable in clinical practice because AKI may occur prior to a steady state. The AUC after the occurrence of AKI is an inappropriate predictor for AKI because high exposure is not the cause of AKI, but the consequence.

In this study, the renal function of six of seven patients with AKI recovered after a reduction in the vancomycin dosage, although they were under a high vancomycin exposure when AKI occurred. The results of causality analyses imply that an elevated exposure of vancomycin is likely to be a risk factor pertinent to AKI occurrence. In addition, neonates may be tolerant to high vancomycin exposure temporarily and vancomycin-induced renal impairments are possibly reversible. The ongoing monitoring of both vancomycin serum concentrations and renal function needs to be emphasised in clinical practice.

Our study reveals that the concomitant use of VAS is not only a significant covariate on vancomycin CL, but also a confounding factor in predicting AKI. Six of seven patients who experienced AKI were co-medicated with VAS. Given that VAS has been reported as an independent risk factor for the development of AKI among ICU neonates[35], vancomycin-induced AKI is probably not only associated with vancomycin exposure alone[2, 8, 9, 36]. Multivariate regression analyses show that high vancomycin exposure (AUC_0-24_ ≥ 485 mg·h/L) remains an independent predictor of AKI after adjusting for the concomitant use of VAS. Although our study cannot explain whether the threshold for AKI will change as a result of concomitant VAS use, there is reason to believe high vancomycin exposure in addition to VAS application contributes to the occurrence of AKI.

The threshold in predicting efficacy in our study (AUC_0-24_/MIC ≥ 234) is far below that in some previous studies in adults (Casapao et al.: AUC_0-24_/MIC ≥ 600; Song et al.: AUC_ss_ over 24 h/MIC ≥ 393)[6, 7]. However, research by Padari et al. showed that an AUC/MIC of ≥ 300 or ≥ 400 does not improve clinical effectiveness in neonates with staphylococcal sepsis[11]. This indicates that a relatively low exposure (AUC/MIC < 300) may still be effective for neonates. In the absence of other efficacy data in neonates to clarify the AUC/MIC necessary for clinical effectiveness against staphylococcal sepsis, the threshold for efficacy needs to be validated in the future.

The relationship between vancomycin exposure and clinical efficacy had not been discovered in previous paediatric and neonatal studies[11, 37]. In contrast, we found that AUC_0-24_/MIC ≥ 234 was strongly correlated with efficacy. This disparity may be caused by several factors. Primarily, the previous study by Hahn et al.[37] focused on patients with various sites of infection, whereas all patients in our study had bloodstream infections. Considering several breakpoints reported by studies on different infectious diseases in adults[4-7], the threshold of vancomycin exposure may be infection site-dependent. Secondly, the whole population of the previous study by Padari et al.[11] had a relatively low vancomycin exposure with only 12% of patients achieving AUC/MIC ≥ 400 compared with 26% in our study. The correlation between efficacy and AUC/MIC may not be detected owing to AUC/MIC values concentrated within a narrow range.

In contrast to the regimen designed in this study, dose recommendations in current referential books are unsuitable for all neonates. The scheme of Neonatal Formulary 7[30] is merely PMA-based, whereas the regimen of Red Book (2018)[31] is based on GA and SCR. According to our Pop-PK model, renal function and the concomitant use of VAS are significant covariates on CL and must be considered in regimen design. Furthermore, regimens in both references[30, 31] are overdosed to ELBW neonates, and the regimen of Red Book (2018)[31] is inadequate for NBW neonates with normal renal function.

There are several limitations to our study. Firstly, our data were derived from a homogeneous neonatal population who had bloodstream infections and were administered vancomycin with intermittent infusion. Therefore, the optimised exposure target should be extrapolated to other populations, who may have different infection sites, with caution. Secondly, given the low morbidity of vancomycin-induced AKI[38] and high effective rate of staphylococcal sepsis in neonates[2, 39], it is important to recognise that the optimised exposure target has limited ability to predict AKI or efficacy. This target can only serve as a guide that balances AKI and efficacy[9]. Finally, the sample size of this study was relatively small. Our findings need to be validated further in large multi-centred prospective studies.

In conclusion, the exposure target of AUC_0-24_ of 240 to 480 (assuming MIC = 1 mg/L) should be advocated to achieve clinical efficacy while minimising side effects for vancomycin treatment in neonates. Model-informed dosing regimens are valuable in clinical practice for ICU neonates with staphylococcal sepsis.

## Data Availability

The data of this study will be open to public through public access of medresman.org.cn within 6 months after publication.

http://www.medresman.org.cn/pub/cn/proj/projectshshow.aspx?proj=1448

## Acknowledgements

We thank Dr Xiaohui Chen and other paediatricians from neonatal ICU of Nanjing Maternity and Child Health Care Hospital. We are also grateful to Dr Xiaojun Ji (Nanjing Chia Tai Tianqing Pharmaceutical Co., Ltd., Nanjing, China) for his assistance with figure preparation.

## Ethical approval

The study was approved by the Ethics Committee of Nanjing Maternity and Child Health Care Hospital (Nanjing, P.R China) according to the Declaration of Helsinki.

## Contributors

Conception and design of the study: Z.T., Z.J.

Acquisition of data: J.G., WW.S., JJ.L., YX.Y.

Analysis of data: Z.T., J.C.

Interpretation of data: Z.T., M.Q.

Drafting and revising the manuscript: Z.T., Z.J.

Revising the manuscript for important intellectual content and final approval of the version for publication: All authors.

## Appendix. Supplementary data

Tables S1 to S9 are available as Supplementary data in the online version.

## Abbreviations^1^

**Abbreviations**
AIC: Akaike information criteria
AKI: acute kidney injury
AUC: area under the concentration–time curve
BIC: Bayesian information criteria
CART: classification and regression tree
CI: confidence interval
CLCR: creatinine clearance
CV: coefficient of variation
cWT: current weight
ELBW: extremely-low-birth-weight
GA: gestational age
ICU: intensive care unit
MAPE: mean absolute prediction error
MC: Monte Carlo
MIC: minimum inhibitory concentration
MPE: mean prediction error
NBW: normal-birth-weight
NPDE: normalised prediction distribution error
NPV: negative predictive value
OFV: objective function value
PMA: postmenstrual age
PNA: postnatal age
Pop-PK/PD: population pharmacokinetic/pharmacodynamic
PPV: positive predictive value
ROC: receiver operating characteristic
SCR: serum creatinine
VAS: vasoactive drug.

